# Economic burden of Respiratory Syncytial Virus Infection in Colombia in 2019: A Cost-of-Illness Study

**DOI:** 10.1101/2025.08.31.25334805

**Authors:** Giancarlo Buitrago, Paula González-Caicedo, Juan Manuel Reyes-Sanchez, Claudia Burgos, Jair Arciniegas, Jorge La Rotta, Omar Escobar, Andreina Alamo

## Abstract

**Background:** Respiratory Syncytial Virus (RSV) is one of the leading causes of acute respiratory infections and severe cases can lead to hospitalization or death. The epidemiology and health resource utilization of RSV infection in Colombia is not well understood. Given the recent availability of new RSV preventatives, this study estimated the economic burden of RSV in Colombia.

**Methods:** This cost-of-illness study employed a retrospective cohort design and bottom-up costing approach to estimate direct healthcare costs associated with RSV-related acute respiratory infections (ARI) across pediatric and adult populations. Administrative data from sentinel surveillance centers belonging to the National Epidemiological Surveillance System of the Colombian National Institute of Health, the database for the study of the Capitation Payment Unit database, and the Integrated Social Protection Information System were utilized to estimate RSV incidence, mortality, and healthcare costs. Costs were expressed in U.S. dollars (USD).

**Results:** A total of 264,744 RSV-related healthcare consultations were identified in 2019. The highest incidence was among infants under one year (61.8 per 1,000), while general mortality was highest in adults ≥75 years (46.6 per 100,000), followed by infants (42.4 per 100,000). Total direct healthcare costs were estimated at $682.87 million (95% CI: $281.39–$1,084.35 million), with the largest share, contributed by individuals aged ≥15 years. Among infants under one year, ICU patients had the highest average cost ($3,619), and hospitalization accounted for 49% of total spending, followed by ICU care (29%) and medications (8%).

**Conclusions:** RSV poses a significant economic burden on Colombia’s healthcare system. These findings support the need for targeted prevention strategies and efficient resource allocation. Future research should incorporate indirect costs and long-term impacts.

**Highlights:**

- The study provides the first national-level estimate of the economic burden of RSV in Colombia using comprehensive administrative data.
- The highest incidence and healthcare costs are observed in infants under one year and adults over 75, highlighting the need for targeted prevention strategies.
- The findings can inform health policy and resource allocation decisions in Colombia’s universal healthcare system.

## BACKGROUND

Respiratory Syncytial Virus (RSV) is one of the leading causes of acute respiratory infections (ARI) worldwide, affecting people across various age groups (1,2). Annually, RSV is responsible for a significant number of hospitalizations and deaths, especially among populations with limited access to healthcare resources (3). It is estimated that more than 20 million RSV episodes occur each year, with approximately 3 million requiring hospitalization, some of which is associated with fatal outcomes (3,4). The highly transmissible nature of RSV places a substantial burden on healthcare systems, highlighting the urgent need for appropriate global prevention and treatment strategies (4).

In recent years, the global distribution of RSV has evolved due to public health measures and social actions, such as those implemented during the coronavirus disease 2019 (COVID-19) pandemic, some of these changes are related to its seasonality, average age of infection and fewer cases during the early phase of the pandemic (5). Despite these measures, RSV remains a major cause of morbidity and mortality in low- and middle-income countries (LMICs), particularly among young children and older adults with comorbidities (3,6). Although RSV is increasingly recognized as a cause of illness in adults, barriers to RSV identification due to limited testing in this population may result in underdiagnosis and a lack of understanding of its true burden (6).

In Colombia, active surveillance of respiratory viruses causing ARI to have been implemented in designated institutions known as sentinel sites. Sentinel units collect respiratory samples from individuals meeting the probable or possible ARI case definitions. These samples are analyzed centrally at the National Institute of Health, and the results guide efforts for control and prevention (7).

Additionally, the behavior of this infection varies depending on factors such as age, region, type of surveillance, case definitions, and diagnostic tests (8,9). However, the magnitude of the RSV infection burden is still limited, representing a gap in knowledge that hinders the development of targeted health policies for controlling this pathogen in the country. Cost-of-illness studies are crucial for evaluating the economic burden of diseases in different countries. These studies justify the implementation of intervention alternatives, support budgetary policy evaluation, and provide a framework for assessing implemented programs (10). In Colombia, cost-of-illness studies have been conducted for chronic non-communicable diseases such as lung cancer and breast cancer (11,12) but are scarce for acute conditions like ARI caused by RSV. Existing RSV studies are single-center analyses focusing solely on pediatric populations, reporting hospitalization costs ranging from US$512 to US$59,501 (13–15).

Colombia’s health system features unique regulatory and coverage aspects that can influence healthcare access and associated costs. The General System of Social Security in Health, established by Law 100 of 1993 (16), outlines three affiliation regimes, covering over 95% of the population under the contributory and subsidized regimes (17). Regardless of the affiliation regime, all individuals are entitled to healthcare services included in a Benefit Plan in Health (PBS), which covers diagnostic and therapeutic services under a cost-effectiveness framework. Health services are provided by Health Promotion Entities, reimbursed through a capitation payment system based on the risk profile of individuals (18). The main objective of this study was to estimate the burden of RSV infection in 2019 as it represents the most recent year prior to the COVID-19 pandemic. This analysis focuses on all PBS-covered costs: medications, hospitalizations, diagnostic and therapeutic procedures, and outpatient services.

## METHODS

This study adopted a cost-of-illness framework using a retrospective cohort design and a bottom-up costing methodology to estimate the burden of acute respiratory infections (ARI) attributable to respiratory syncytial virus (RSV) in Colombia during 2019. The analysis was conducted from the perspective of the healthcare system as a third-party payer, encompassing both pediatric and adult populations diagnosed with RSV. The number of RSV-related cases was first estimated, followed by the calculation of direct healthcare costs based on individual-level service utilization.

All cost data reflect direct medical expenditures billed and reimbursed within the Colombian health system for services included in the national Benefit Plan in Health (PBS). Costs originally recorded in Colombian pesos (COP) for the year 2019 were adjusted for inflation using the official Colombian consumer price index and subsequently converted to U.S. dollars (USD) at the representative exchange rate as of December 31, 2023 (1 USD = 3,822 COP) (19). The analysis excluded non-reimbursed costs such as home care, patient transportation, out-of-pocket payments, and productivity losses.

### Data Sources

This study drew upon multiple administrative data sources from the Colombian healthcare system, including publicly available datasets and databases provided by the Ministry of Health and Social Protection to the Institute of Clinical Research at the National University of Colombia for research purposes. The databases from government entities were delivered duly anonymized, ensuring that individual patients could not be identified during the study; they were accessed in the period comprehended between 1-Feb-2024 and 30-Nov-2024. The primary source was a dataset derived from four selected sentinel surveillance centers, which are part of the National Public Health Surveillance System (SIVIGILA), coordinated by the Colombian National Institute of Health (Instituto Nacional de Salud, INS). These centers conduct ongoing surveillance of ARI, systematically collecting respiratory samples from individuals who meet the probable or possible case definitions. RSV infections are confirmed through polymerase chain reaction (PCR) testing conducted in national reference laboratories (7,20). The anonymized dataset includes variables such as birth date, dates of symptom onset and healthcare consultation (inpatient and outpatient), sampling site, and laboratory-confirmed test results for all suspected ARI cases identified through this sentinel system.

The second source was the database from the sufficiency study of the Capitation Payment Unit (*Unidad de Pago por Capitación* - UPC), utilized by Colombia’s Ministry of Health and Social Protection to estimate the annual capitation premium provided to each Health Promotion Entity (*Entidad Promotora de Salud* - EPS). This premium covers the projected healthcare service expenses for each EPS member in the subsequent year. The database contains detailed and standardized information on every healthcare service or technology utilized within the contributory health regime during a calendar year, including the date of service provision, the corresponding ICD-10 diagnostic code, service or medication codes, the healthcare provider delivering the service or medication, the municipality, and the payment made by the insurer to the provider. The database exclusively includes data from insurers meeting rigorous validation standards set by the Ministry, representing approximately 85% of all patients affiliated with the contributory regime in 2019. Further details on this database can be found in Bolívar et al. (21).

To approximate the number of RSV cases nationwide, a third source, the Integrated Social Protection Information System (SISPRO) data cubes, was used. These multidimensional structures organize health affiliation statistics, allowing data exploration by sex, age group, geographical region, and type of health affiliation (contributory/subsidized) (22). Mortality data were obtained from the Global Burden of Disease (GBD) summaries, detailing the number of deaths from various conditions, including lower respiratory infections associated with RSV (23).

### Patient Identification and Case Estimation

To estimate the total number of RSV-related acute respiratory infection (ARI) cases in both contributory and subsidized health regimes, we initially used confirmed RSV-positive ARI case data from sentinel surveillance centers affiliated with the contributory regime. Using the UPC database, we identified all suspected ARI cases reported by these sentinel centers and calculated the proportion of RSV-positive cases among these suspected ARI patients, stratified by age group. Next, we determined the total number of suspected ARI cases nationwide within the contributory regime using detailed microdata available only for this regime. Dividing this number by the affiliated population within each age category represented in the UPC database allowed us to estimate the proportion of contributory regime members consulting healthcare services due to suspected ARI. These proportions were then extrapolated to the subsidized regime under the assumption that healthcare utilization patterns and RSV positivity proportions were similar between both regimes. Finally, by applying the calculated RSV positivity proportions to the estimated suspected ARI cases, we determined the total number of probable RSV-related ARI cases seeking healthcare services in Colombia in 2019, separately for each health regime.

### Incidence and Mortality Estimation

The incidence rate of RSV in Colombia was calculated as the ratio of new RSV cases (estimated) to the total at-risk population during the study period multiplied by 100,000, based on the number of affiliates reported in SISPRO for the contributory regime in 2019:

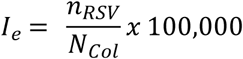

Where 𝐼_𝑒_ is the incidence rate in each category; 𝑛_𝑅𝑆𝑉_ is the estimated number of RSV cases; and 𝑁_𝐶𝑜𝑙_ is the At-risk population in Colombia.

For mortality estimation, the ratio of new RSV-related deaths to the total population affiliated with the health regimes, as reported in SISPRO for the study period, was calculated using GBD data about mortality, extrapolating it to the Colombian population with SISPRO data. Incidence and mortality estimates were made for individuals in the subsidized scheme to provide a comprehensive national perspective.

The full process for estimating RSV cases, incidence, and mortality is shown in Figure 1.

**Figure 1.**
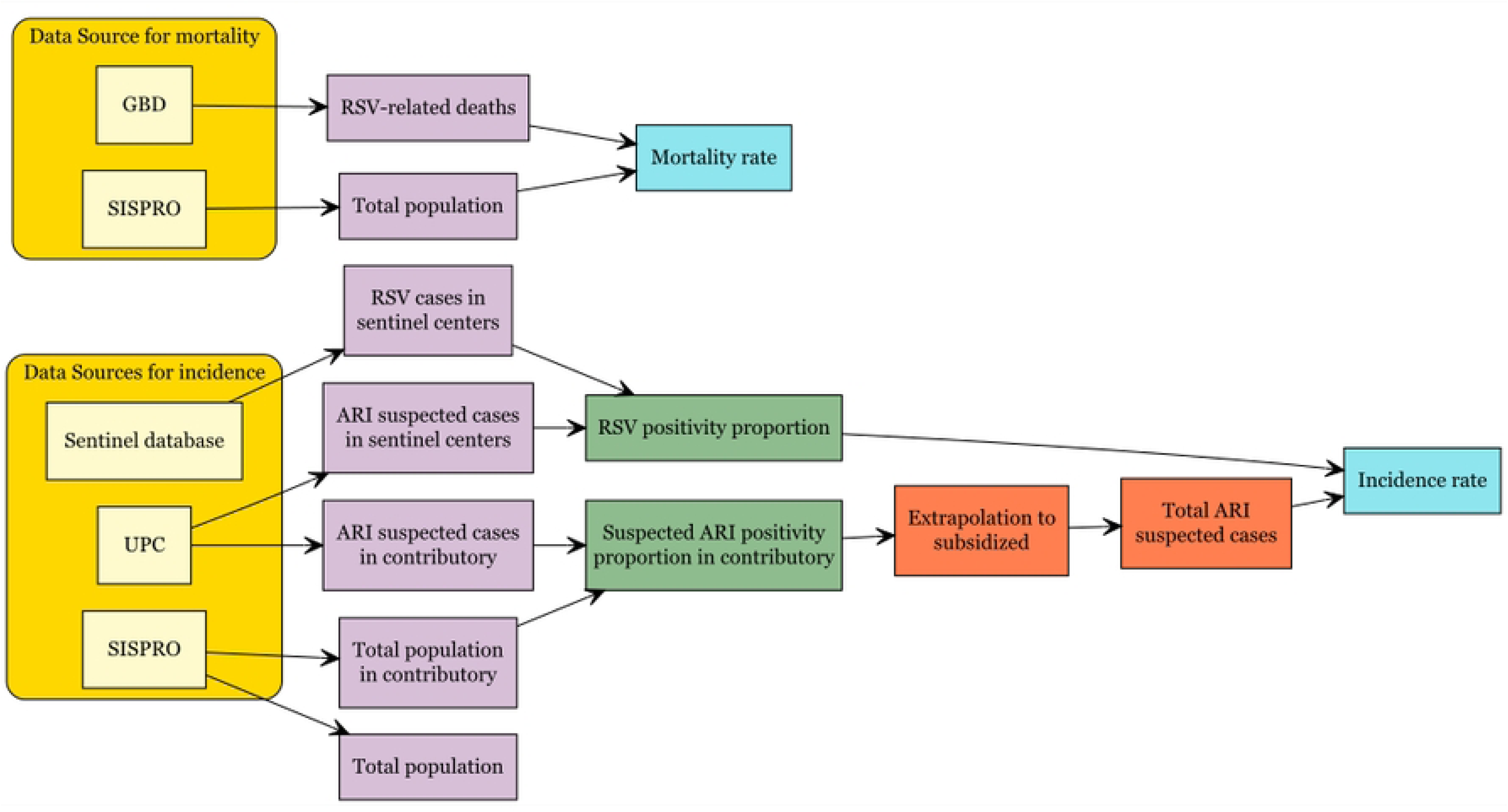
Estimation methods. Approach for the estimation of incidence and mortality associated with RSV infections in Colombia (2019).

### Cost Estimation at 30 Days

Direct healthcare costs were estimated using individual-level claims data from the UPC database, capturing all billed services provided from the date of RSV diagnosis (cohort entry) through either death or the end of a 30-day follow-up period. Cost estimates included all services reimbursed by the health system under the Benefit Plan in Health (PBS) and were stratified by patient age group and healthcare setting.

For infants under one year of age—a group with the highest incidence rates—costs were further disaggregated by the highest level of care received during the follow-up period: outpatient care, general hospitalization, or intensive care unit (ICU) admission. In this classification, each patient was assigned to a single category based on the most intensive setting used, and total costs included all services received, not just those provided within that setting.

In addition, a component-based analysis was conducted to quantify the proportion of total healthcare expenditures attributable to major service categories—namely, hospitalizations, ICU stays, medications, medical consultations, and other services (including laboratory tests, diagnostic imaging, rehabilitation, patient transfers, and emergency department observation). This allowed for a detailed assessment of cost drivers, regardless of the healthcare setting in which the patient was treated.

All analyses were performed using Stata MP® 17.0 and Microsoft® Excel® for Microsoft 365 MSO, under a license from the Universidad Nacional de Colombia.

## RESULTS

### Identification of RSV-Infected Patients Who Sought Healthcare Services

The RSV positivity proportion and ARI positivity proportion identified in sentinel centers are reported in Supplementary Table 1. After extrapolating suspected ARI patients from contributory regime with the proportions previously mentioned, 264,744 RSV-ARI infection cases in Colombia who sought healthcare services during 2019 were identified, including contributory and subsidized regimes. The estimate was highest among the population aged 15 to 49 years (38.62% [n = 102,236]) due to the concentration of Colombian population in that age group, also it was higher in individuals affiliated with the subsidized health regime (53.24% [n = 140,951]). Table 1 presents the detailed distribution by age groups and health affiliation regime.

**Table 1.**
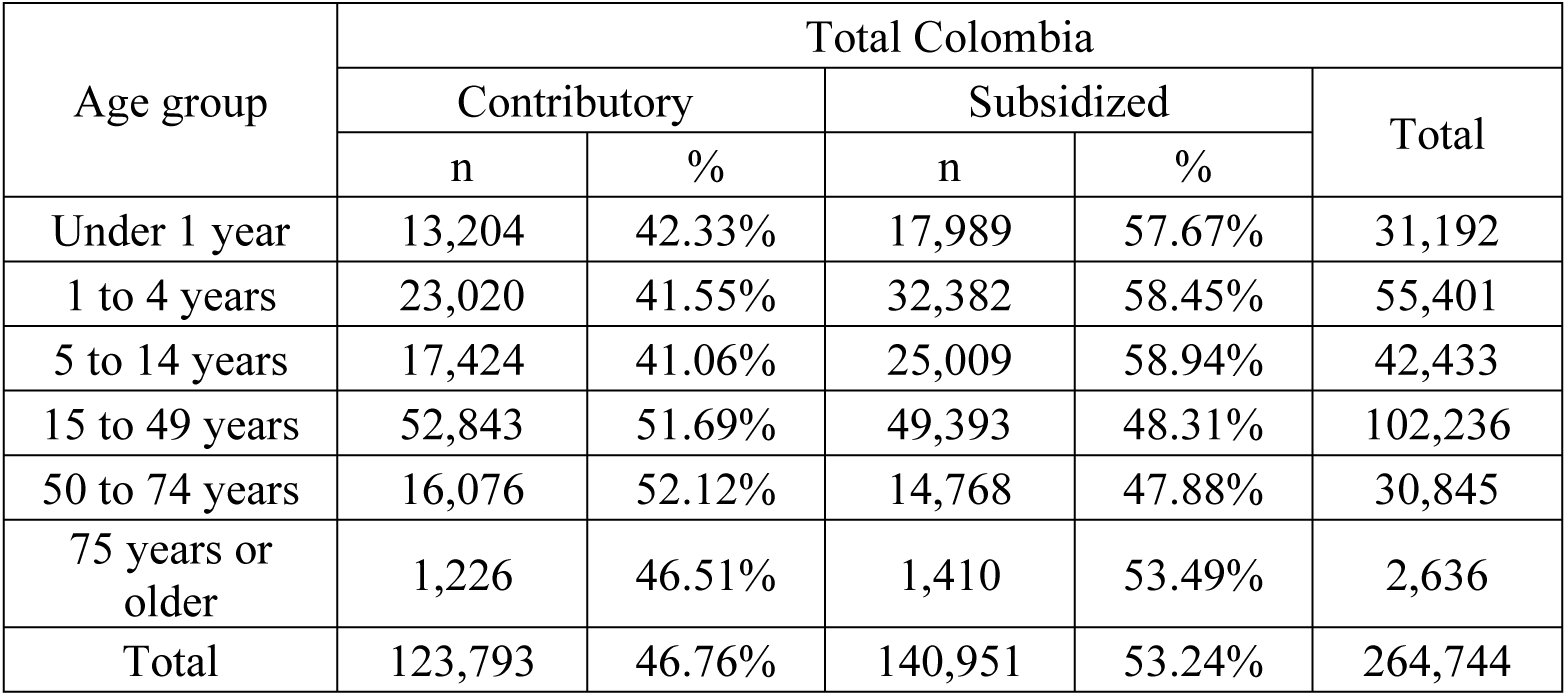
Estimated number of patients with Respiratory Syncytial Virus (RSV) infection who sought healthcare services in Colombia (2019), stratified by age group and health insurance regime.

### RSV-Related ARI Incidence

The incidence of RSV in patients who sought healthcare services in Colombia in 2019 was 582 cases per 100,000 individuals. The estimate was highest among infants under one year (6,180 cases per 100,000), followed by children aged 1 to 4 years (2,221 cases per 100,000) (see Figure 2). Differences were also observed between health regimes: 618 cases per 100,000 individuals in the subsidized regime compared to 546 cases per 100,000 in the contributory regime.

**Figure 2.**
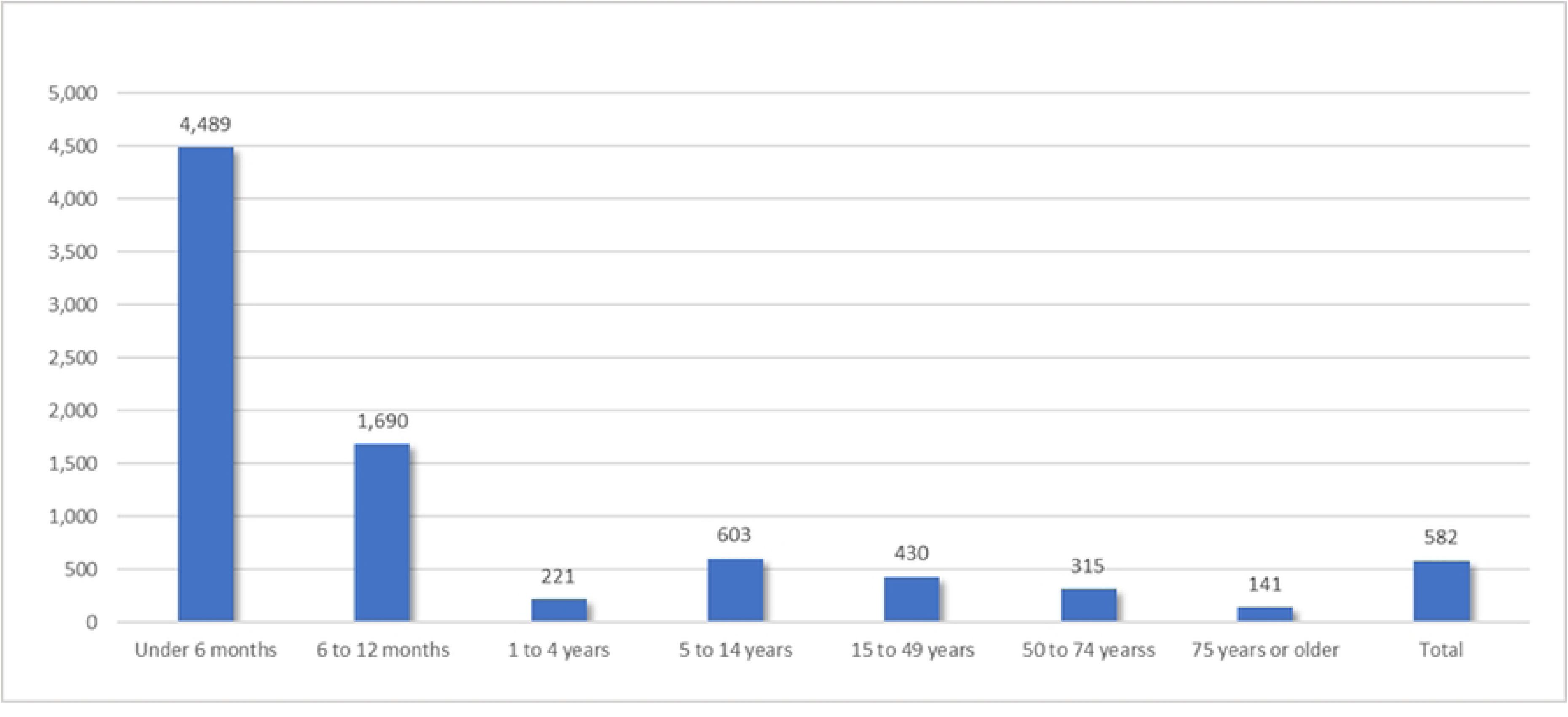
Incidence of RSV infection. Estimated incidence rate (per 100,000 insured individuals) of RSV infections among patients who sought healthcare services in Colombia (2019), stratified by age group.

An analysis by healthcare setting revealed a higher incidence in hospitalized patients than in outpatient or emergency settings (643,80 vs. 56,50 and 34,24 per 100,000 affiliates, respectively); this pattern was consistent across all age groups (see Table 2).

**Table 2.** Estimated incidence rates (per 100,000 insured individuals) of RSV infections in Colombia (2019), by age group, health insurance regime, and type of healthcare service (hospitalization, emergency department, and outpatient care).

| Age group | Contributory |  |  | Subsidized |  |  | Total Colombia |  |  |
| --- | --- | --- | --- | --- | --- | --- | --- | --- | --- |
|  | Hospitalization | Emergency Department | Outpatient | Hospitalization | Emergency Department | Outpatient | Hospitalization | Emergency Department | Outpatient |
| Under 6 months | 2996.70 | 211.22 | 52.81 | 2997.61 | 211.29 | 52.82 | 2997.23 | 211.26 | 52.81 |
| 6 to 12 months | 437.41 | 14.91 | 9.94 | 437.54 | 14.92 | 9.94 | 437.48 | 14.91 | 9.94 |
| 1 to 4 years | 293.97 | 71.86 | 58.79 | 293.95 | 71.86 | 58.79 | 293.96 | 71.86 | 58.79 |
| 5 to 14 years | 7.09 | 1.77 | 1.77 | 7.09 | 1.77 | 1.77 | 7.09 | 1.77 | 1.77 |
| 15 to 49 years | 4.92 | 1.23 | 1.23 | 5.21 | 1.30 | 1.30 | 5.06 | 1.26 | 1.26 |
| 50 to 74 years | 6.57 | 0.94 | 1.88 | 6.41 | 0.92 | 1.83 | 6.49 | 0.93 | 1.86 |
| 75 years or older | 0.42 | 0.00 | 0.42 | 0.41 | 0.00 | 0.41 | 0.41 | 0.00 | 0.41 |
| Total | 603.95 | 53.01 | 32.13 | 683.40 | 59.98 | 36.35 | 643.80 | 56.50 | 34.24 |

### RSV-Related ARI Mortality

Cumulative mortality from RSV in 2019 was 3.37 per 100,000 individuals, similar in those affiliated with the contributory regime compared with the subsidized regime (6.76 vs 6.72 per 100,000, respectively; see Figure 3). By age group, the highest mortality rates were observed in individuals aged 75 years and older (46.55 per 100,000) and in infants under one year (42.40 per 100,000). The number of deaths are reported in Supplementary Table 2.

**Figure 3.**
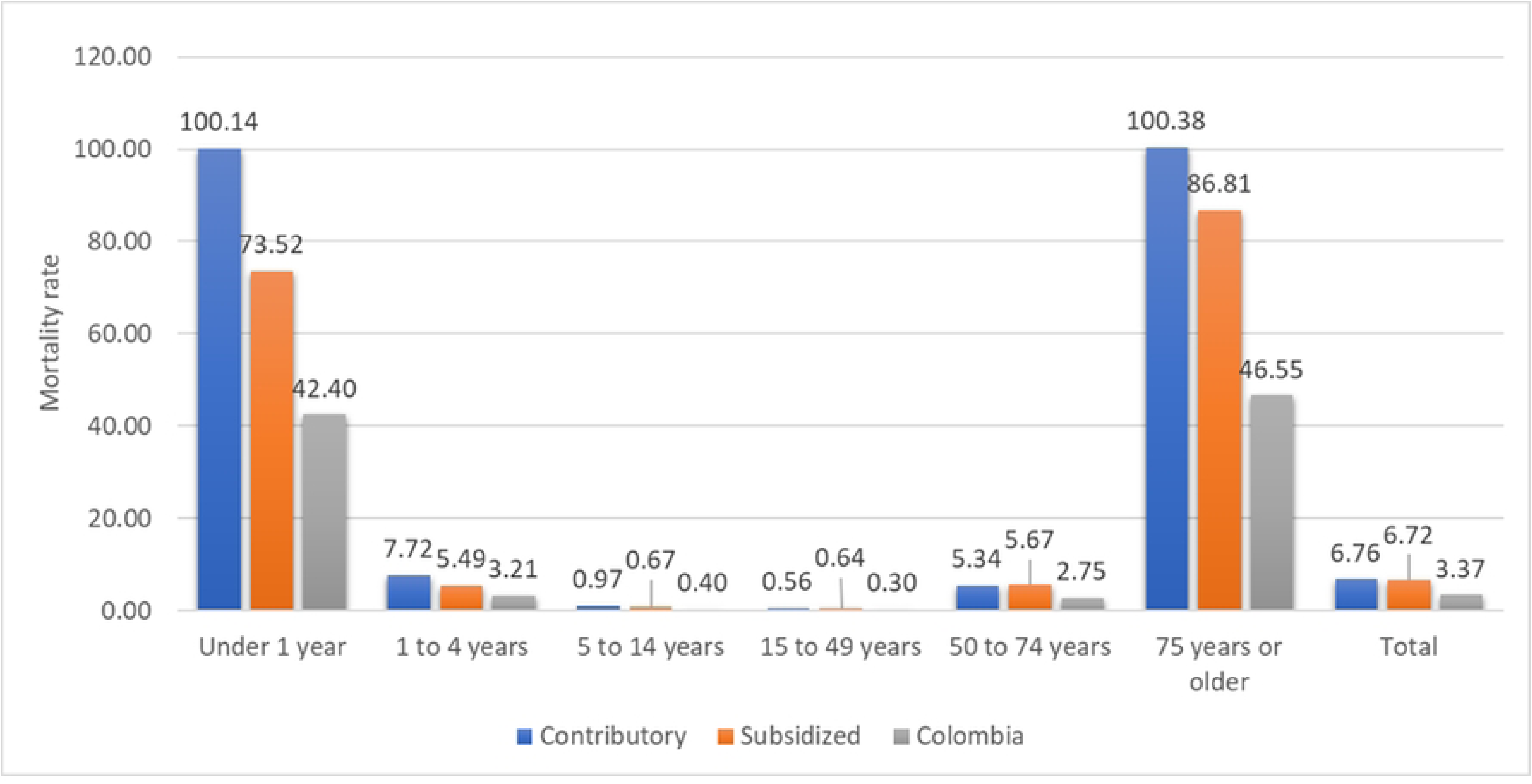
Mortality related to RSV infection. Mortality rates (per 100,000 insured individuals) associated with RSV infection among patients who sought healthcare services in Colombia (2019), stratified by age group.

### Cost Estimation

The total direct healthcare costs associated with RSV-related healthcare services in Colombia in 2019 were estimated at $682.87 million (95% CI: $281.39 – $1,084.35 million) (see Table 3). Total costs observed among individuals aged 15 years and older were $480.40 million (95% CI: $158.94 – $801.86 million). The total for infants under one year was $52.32 million (95% CI: $44.59 – $60.05 million).

**Table 3.** Total and per-patient direct healthcare costs associated with RSV infection in Colombia (2019), by age group.

| Age group | Total Costs in million* (95% CI) | Costs per patient (95% CI) |
| --- | --- | --- |
| Under 1 year | 52.32 (44.59 - 60.05) | 1,677 (1,430 – 1,925) |
| 1 to 14 years | 150.15 (77.85 - 222.44) | 1,535 (796 – 2,274) |
| 15 years or older | 480.40 (158.94 - 801.86) | 3,540 (1,171 – 5,908) |
| Total | 682.87 (281.39 – 1 084.35) | 2,579 (1,063 – 4,096) |

\*USD in 2019

When examining average costs per patient, the overall mean was estimated at $2,579 (95% CI: $1,063 – $4,096). The highest per-patient costs occurred among individuals aged 15 years or older ($3,540; 95% CI: $1,171 – $5,908), reflecting more severe clinical presentations and intensive resource utilization within this age group. The second highest costs were found in children under 1 year, whose average cost was ($1,677; 95% CI: $1,430 – $1,925). Conversely, the lowest average cost per patient was identified in children aged 1 to 14 years ($1,535; 95% CI: $796 – $2,274). Table 3 presents detailed cost estimations by age group.

A detailed cost analysis for infants under one year of age (Table 4) revealed that intensive care unit (ICU) services represented the highest average cost per patient, at $3,619 (95% CI: $2,750 – $4,489), accounting for approximately half of the total healthcare expenditures ($24.57 million) in this age group. Hospitalizations contributed the largest absolute share of total costs, amounting to $26.19 million (95% CI: $22.52 – $29.87 million), with an average cost per patient of $1,225 (95% CI: $1,053 – $1,397). Outpatient care represented the smallest proportion of costs, totaling $1.56 million (95% CI: $0.77 – $2.34 million), with a mean cost per patient of $514 (95% CI: $253 – $774). In all healthcare service categories, costs were consistently lower for infants aged 6 to 12 months compared to those younger than 6 months.

**Table 4.** Total and per-patient direct healthcare costs among children under one year with RSV infection in Colombia (2019), by type of healthcare setting.

| Care setting | Total costs in million* (95% CI) |  |  | Cost per patient* (95% CI) |  |  |
| --- | --- | --- | --- | --- | --- | --- |
|  | Under 6 months | 6 to 12 months | Total | Under 6 months | 6 to 12 months | Total |
| Outpatient | 1.42<br>(0.64 – 2.19) | 0.14<br>(*** – 0.31) | 1.56<br>(0.77 – 2.34) | 572<br>(259 – 885) | 249<br>(** – 564) | 514 (253 – 774) |
| Hospitalized | 19.75<br>(16.35 – 23.16) | 6.44<br>(5.08 – 7.80) | 26.19<br>(22.52 – 29.87) | 1,329<br>(1,100 – 1,558) | 988<br>(780 – 1,197) | 1,225<br>(1,053 – 1,397) |
| ICU | 18.37<br>(14.30 – 22.45) | 6.20<br>(1.52 – 10.87) | 24.57<br>(18.67 – 30.47) | 3,453<br>(2,687 – 4,220) | 4,223<br>(1,038 – 7,408) | 3,619<br>(2,750 – 4,489) |
| Total | 39.55<br>(33.58 – 45.51) | 12.77<br>(7.80 – 17.75) | 52.32<br>(44.59 – 60.05) | 1,745<br>(1,482 – 2,009) | 1,497<br>(0,914 – 2,080) | 1,677<br>(1,430 – 1,925) |

\*Cost in 2019 USD
\*\* Not estimable

Furthermore, the distribution of healthcare costs by type of service (Supplementary Figure 1) indicated that hospital admissions accounted for nearly half (49.0%) of the total expenditures among infants under one year, followed by ICU admissions (28.6%). Medications represented 7.9%, outpatient consultations 5.0%, and other healthcare services—including laboratory tests, diagnostic imaging, rehabilitation, patient transfers, and emergency department observations—collectively accounted for 9.5% of total expenditures.

A more granular analysis comparing infants under 6 months to those aged 6 to 12 months showed a similar overall distribution of costs by service type. However, notable differences emerged in the proportion of costs attributable to ICU admissions, which was higher in younger infants (30.6%) than in older infants (22.2%). Conversely, expenditure on medications was proportionally higher for infants aged 6 to 12 months (12.3%) compared to those under 6 months (6.5%).

## DISCUSSION

This study estimated the burden of RSV infection in 2019 for Colombia, a middle-income country with universal healthcare coverage. The analysis was stratified by age group, and health affiliation regime (contributory and subsidized). To our knowledge, this is the first study to use this approach to report a national estimate of the RSV burden. The findings of this study are consistent with those presented by Shi et al. (24), who estimated the global number of ARI episodes caused by RSV, reporting that the highest proportion of cases requiring hospital treatment occurs among infants under one year of age. This is likely due to the immaturity of the immune system and a higher risk of respiratory complications due to increased airway resistance, higher metabolic oxygen demand, inefficient respiratory muscles and small functional residual capacity, compared to adults. (25)

For children under five, the incidence in this study aligns with Hall et al. (26), who found higher incidence rates among infants under one year compared to older pediatric age groups. This underscores the need to prioritize prevention in infants under one year as they are highly susceptible to severe complications and death during surge of respiratory infections (27). For adults aged 75 and older, our findings are also consistent with studies highlighting the vulnerability of this population to RSV, particularly those with comorbidities. While the incidence among older adults is lower than in pediatric populations in the current study, RSV can still be a significant cause of morbidity and mortality as demonstrated in our analysis and in several studies (28,29). Recent publication evaluating RSV hospitalization rates in Argentina, Brazil, Chile, and Mexico using a modeling approach estimated rates of 132.8 to 816.5 per 100,000 in adults aged ≥75 years, far greater than what is reported in the current study (30). Additional work should be undertaken to better understand the burden in older adults and adults with comorbidities as many limitations apply in identifying cases of RSV in adults including challenges in diagnostic sensitivity (31–33).

Our study estimated an incidence rate of 61.80 per 1,000 children under one year, similar to the 63.3 per 1,000 reported by Shi et al., based on a meta-analysis of 329 studies estimating the RSV burden in LMICs (24). Our findings on the estimated mortality rate of 3.37 per 100,000 are comparable to those reported by Du et al., who documented a global mortality rate of 4.79 per 100,000 individuals. The study found that RSV mortality rates decline and then rise with age, with higher rates among those over 70 years compared to children under five. This pattern is consistent with our findings, although we report higher mortality rates among infants under one year (27). Gómez et al. (34) estimated RSV-associated hospitalization and mortality burden in adults in five middle-income countries, finding trends similar to ours in four countries (excluding Argentina). This reinforces previous research showing that both young children and the elderly are at higher risk of severe RSV outcomes (26,28,29,35,36).

Our cost analysis revealed a total amount of $682.87 million (95% CI: $281.39 million – $1,084.358 million), or an average cost per patient is $2,579 thousand (95% CI: $1,063 – $4,096) indicating a substantial economic burden. The costs found in the study are consistent with previous studies, such as Rodríguez-Martínez et al. (37), which reported per-patient hospitalization costs in Colombia ranging from $518 in pediatric wards to $2,749 in intensive care units. Buendía et al. (13) also conducted a cost-of-illness study for RSV in infants from Colombia, estimating an annual total cost of $64.44 million (95% CI: $11.09 – $195.72 million) in children under two years. Significant economic burden associated with infant RSV infection was also reported outside of Colombia. Total annual hospitalization costs for infants < 1 year totaled 30.9 MM euros (see figure 1, adding 0 - 5 m and 6 - 11 m for RSV-specific hospitalizations costs) (38). There is a comprehensive approach that includes outpatient and emergency care, offering a broader economic impact perspective.

While the epidemiologic burden of RSV in older adults and adults with comorbidities are increasingly being recognized, the economic burden reported from Latin America region is limited. A study in Argentina reported a mean hospital length of stay of 12 days with 37% requiring intensive care unit admission in adults with confirmed RSV infection, but no costs were reported (39). Based on national data in Spain, a mean cost of RSV hospitalization of €3,870 in adults and a total RSV-attributable hospitalization cost per year of €194 million was recently reported (40). Additional studies in Colombia and Latin America region are needed to better characterize the economic burden of RSV in adults.

This study has several strengths. First, it employs a bottom-up cost-of-illness approach based on individual-level administrative data, enabling a detailed and accurate estimation of the RSV burden from the healthcare system perspective. Second, it provides a comprehensive national analysis across all age groups and both major health insurance regimes (contributory and subsidized), which is rarely addressed in previous studies. Third, unlike prior research that focused primarily on hospitalization costs, this study incorporates costs associated with outpatient, emergency, and intensive care services, offering a broader and more realistic picture of the economic impact of RSV in Colombia.

Nonetheless, some limitations must be acknowledged. The analysis required extrapolation from the contributory regime to the subsidized population due to the lack of individual-level cost data for the latter, which may introduce bias if healthcare-seeking behavior or disease severity differs across regimes. Additionally, the cost analysis was restricted to services covered under the Benefit Plan in Health (PBS), excluding indirect costs such as transportation, productivity losses, and out-of-pocket expenditures, potentially underestimating the total economic burden. Furthermore, the study period precedes the COVID-19 pandemic; thus, results may not reflect current RSV epidemiology or healthcare system dynamics in the post-pandemic context. It is also worth noting that the costs presented in the study are all-cause healthcare utilization and not specific to RSV-related treatment, resulting in a potential overestimation of the costs; however, they are not expected to differ significantly.

Another limitation is the potential under ascertainment of RSV cases, specifically in the adult population. Although PCR testing is systematically performed for all suspected ARI cases in sentinel hospitals, diagnostic accuracy—particularly in adults—is challenged by the lower sensitivity of nasopharyngeal swabs and the need for complementary respiratory specimens or serological tests, as such, case detection is incomplete (31–33). According to recent systematic literature review and modeling studies, the true magnitude of the RSV hospitalization could be 2.2 times what is reported in surveillance studies (41). Moreover, sentinel hospitals only capture patients who seek healthcare services; this causes an exclusion of milder cases, which are more common in this population, and results in potential selection bias. According to Chavez et al. (42), in Bolivia the mean incidence of RSV in adult population from the period 2013-2016 was 77.4 cases for adults aged between 20 and 64 and 607.5 for adults aged 65 and older. The findings from the literature confirm that the RSV incidence rates in adults from the current study are underestimated.

## CONCLUSIONS

This study identified a significant disease burden of RSV infection in Colombia, affecting mainly pediatric but also older adult populations. These estimates may inform public health policies aimed at optimizing health outcomes in the Colombian population and guide resource allocation decisions. Future research should consider both direct healthcare costs and indirect and intangible costs associated with the long-term consequences of RSV-related ARI. Our findings emphasize the need for primary prevention strategies and further research to identify mechanisms for optimizing the observed RSV-related costs in Colombia, addressing broader impacts on productivity, caregiver burden, and education. Effective public health interventions could alleviate the substantial morbidity, mortality, and economic burden imposed by RSV, particularly in middle-income countries like Colombia.

## DECLARATIONS

### Patient consent for publication

Not applicable.

### Ethics approval

This study was approved by the Ethics Committee of the Faculty of Medicine, Universidad Nacional de Colombia (Act No. 018, October 26, 2023).

### Availability of data and materials

The following information sources: Unique Affiliation Database (Base de Datos Única de Afiliación, or BDUA), and Calculation Study of the Capitation Unit Database (Base del Estudio de Suficiencia de la Unidad Por Capitación, or UPC) are administered by the Colombian Ministry of Health and Social Protection. These databases are freely available upon request to the Technology of the Information and Communication Office of the Colombian Ministry of Health and Social Protection through the.

### Authors’ contributions

Conceptualization: GB. Data collection: GB, PG-C. Formal analysis: GB, PG-C. Methodology: GB, JR, JA. Investigation: GB, PG-C, JR, AA, JA. Project administration: JR, AA. Software: GB, PG-C. Supervision: JR, AA, OE, JLR. Validation: AA, CB. Visualization: GB, PG-C. Writing original draft: GB, PG-C, AA. Writing review and editing: GB, AA, CB, OE, JLR. The authors read and gave final approval of the manuscript.

## ACKNOWLEDGMENTS

We thank Colombia’s Ministry of Health and Social Protection for providing the administrative databases that made this study possible. We also thank the Clinical Research Student Group at the Faculty of Medicine of the National University of Colombia for their valuable contributions. This study’s preliminary findings were presented and discussed within this group, enriching the interpretation and scope of our results. The School of Medicine at Universidad Nacional de Colombia supported the study.

## ABBREVIATIONS

RSV: Respiratory Syncytial Virus.
USD: United Sates Dollars.
ARI: Acute Respiratory Infections.
SIVIGILA: National Epidemiological Surveillance System.
INS: Colombian National Institute of Health.
UPC: Capitation Payment Unit.
SISPRO: Integrated Social Protection Information System.
LMICs: low- and middle-income countries.
PBS: Benefit Plan in Health.
COP: Colombian pesos.
PCR: Polymerase chain reaction.
EPS: Health Maintenance Organisation.
GBD: Global Burden of Disease.
ICU: Intensive Care Unit.
BDUA: Unique Affiliation Database.

## Notes

### Competing Interest Statement

I have read the journal's policy and the authors of this manuscript listed below have the following competing interests: AA, JA, JR, JLR and CB are paid employees of Pfizer.

